# Online social networks of individuals with adverse childhood experiences

**DOI:** 10.1101/2022.12.19.22283651

**Authors:** Yiding Cao, Suraj Rajendran, Prathic Sundararajan, Royal Law, Sarah Bacon, Steven A. Sumner, Naoki Masuda

**Affiliations:** Department of Systems Science and Industrial Engineering, Binghamton University, State University of New York, Binghamton, NY 13902-6000, USA; Tri-Institutional Program in Computational Biology and Medicine, New York, NY, USA; Department of Biomedical Engineering, Georgia Institute of Technology, Atlanta, GA, USA; Division of Injury Prevention, National Center for Injury Prevention and Control, Centers for Disease Control and Prevention. Atlanta, GA, USA; Office of Strategy and Innovation, National Center for Injury Prevention and Control, Centers for Disease Control and Prevention. Atlanta, GA, USA; Department of Mathematics, State University of New York at Buffalo, Buffalo, NY, USA; Computational and Data-Enabled Science and Engineering Program, State University of New York at Buffalo, Buffalo, NY, USA

**Author notes:** Equal contribution (Suraj Rajendran, Prathic Sundararajan).

## Abstract

Adverse childhood experiences (ACEs), which include abuse and neglect and various household challenges like exposure to intimate partner violence and substance use in the home can have negative impacts on lifelong health of affected individuals. Among various strategies for mitigating the adverse effects of ACEs is to enhance connectedness and social support for those who have experienced ACEs. However, how social networks of those who experienced ACEs differ from those who did not is poorly understood. In the present study, we use Reddit and Twitter data to investigate and compare social networks among individuals with and without ACEs exposure. We first use a neural network classifier to identify the presence or absence of public ACEs disclosures in social media posts. We then analyze egocentric social networks comparing individuals with self-reported ACEs to those with no reported history. We found that, although individuals reporting ACEs had fewer total followers in online social networks, they had higher reciprocity in following behavior (i.e., mutual following with other users), a higher tendency to follow and be followed by other individuals with ACEs, and a higher tendency to follow back individuals with ACEs rather than individuals without ACEs. These results imply that individuals with ACEs may try to actively connect to others having similar prior traumatic experiences as a positive connection and coping strategy. Supportive interpersonal connections online for individuals with ACEs appear to be a prevalent behavior and may be a way to enhance social connectedness and resilience in those who have experienced ACEs.

## 1 Introduction

Adverse Childhood Experiences, or ACEs, are preventable, potentially traumatic events that occur in childhood (0-17 years) such as being neglected, experiencing or witnessing violence, and having a family member attempt or die by suicide. Also included are aspects of a child’s environment that can undermine their sense of safety, stability, and bonding, such as growing up in a household with substance use, mental health problems, or instability due to parental separation or incarceration of a parent, sibling, or other member of the household [1, 2]. These examples do not comprise an exhaustive list of childhood adversity, as there are other traumatic experiences that could impact health and wellbeing. ACEs often occur together, can result in toxic stress, and are associated with a wide range of adverse behavioral, health, and social outcomes, including substance use, depression, overweight/obesity, lower education and earnings potential, and chronic diseases such as heart disease and cancer. ACEs are preventable. Prevalence of ACEs is estimated to be as high as 60% in the United States [3, 4, 5] and worldwide [4]. With the advent of the COVID-19 pandemic in early 2020 and the following economic stress and instability within the United States, there has been an increase of stress in parenting [6], which may contribute to increased risk for ACEs [7, 8].

Connecting youth to caring adults and activities is an evidence-based strategy for the primary prevention of ACEs. An extension of this strategy is that connectedness and social support have been suggested to play a significant role in mitigating negative impacts of ACEs across the lifespan. In fact, longitudinal studies have shown that individuals with ACEs tend to have less interpersonal social support than those without ACEs when they became adults [9–11]. Although social support as a mitigating factor in various health problems has been studied for decades, the definition of social support can vary [12, 13]. It has been suggested that social networks of individuals, i.e., how they are connected to each other through social ties and embedded in social groups, can be a powerful form of social support [14, 15]. Understanding the characteristics of social networks of individuals who discuss their ACEs and their health are useful for identifying where and how social support can be improved.

Studies of social networks of individuals reporting ACEs exposure have largely focused on examining the numbers and types of other individuals in close physical proximity with whom they directly interact and the quality of such social ties [16, 17]. However, social networks are not only defined by numbers and proximity but also defined by the relationship between individuals. Particularly in the modern, internet-enabled culture, individuals are embedded in a larger social network through which information and support can flow in more expansive ways than previously possible [18, 19]. Social media is universally used to discuss various topics including health, and ACEs are no exception [20]. Social media information has also been used to estimate mental health status of individuals [21–24]. These and other results suggest opportunities for researchers to use social media information to better understand conversations about ACEs, understand health-related information pertaining to ACEs, and enhance social support among those with ACEs. Prior research in other health topics including major depression [25, 26, 27], suicide ideation [25, 28], anxiety disorder [29], and schizophrenia [30] have revealed that characteristics of an individual’s online social network bears an association with aspects of health status.

Motivated by this, we use data from two social media platforms to examine the online social network of individuals reporting ACEs and discuss implications for public health.

## 2 Methods

This work involved a multi-step pipeline to first create a classifier to identify ACEs content from social media posts, deploy the classifier to a large body of social media posts, and compute network statistics from such information.

### 2.1 Reddit data

To first construct a machine learning based, automated classifier for self-reported ACEs disclosures, we used a transfer learning approach commonly used in online data research [31]. Using the Pushshift.io Reddit API (https://github.com/pushshift/api), we downloaded Reddit data from two subreddits: r/raisedbynarcissists and r/internetparents. Subreddit r/raisedbynarcissists is a support group for people raised by abusive parents and hence contains many posts detailing experiences of ACEs. Subreddit r/internetparents is structured similarly to r/raisedbynarcissists but focuses on generally positive childhood events and experiences. We use these two subreddits as the explicit labels to build a binary supervised learning classifier with r/raisedbynarcissists as the positive class, associated with the presence of ACEs, and r/internetparents as the negative class, associated with the absence of ACEs. Note that we could not use Twitter data for training a classifier because tweets do not have ACE-related labels. We collected all available posts within the two subreddits between December 25, 2020, and March 31, 2022, totaling 49,044 posts from r/raisedbynarcissists and 21,712 posts from r/internetparents. We used the title of the post to train the classifier because our investigation revealed that post titles are similar in length to tweets and contain sufficient information about the ACE experienced.

Before submitting the Reddit post titles to the training of the classifier, we cleaned the post titles from both subreddits using the Python-based regex and Natural Language Toolkit (NLTK) (https://www.nltk.org/) libraries. Our NLP pipeline to clean text included dropping duplicate post titles, expanding abbreviations, and removing special characters, among others. We deleted all post titles with ≤ 5 words to improve training of the classifier. There were in total 22,950 posts (70%) from r/raisedbynarcissists and 9,744 posts (30%) from r/internetparents that were ultimately used in the model. Details on the text preprocessing are available in Github (https://github.com/ycao20/ACE-project).

### 2.2 Convolutional neural network classifier

We used a convolutional neural network (CNN) for our classifier; CNNs have successfully been applied to image and text processing [32, 33]. CNNs have also been employed in detecting mental health conditions from Reddit data [34, 35]. CNNs work by modeling hierarchical complicated patterns using smaller and simpler patterns. Convolutional layers along with the max-pooling layer allows the CNNs to learn useful word representations while enhancing their computational efficiency.

We trained a CNN using the Reddit post title as input and the class label as teacher. Because there are more data in the positive class than in the negative class, we selected samples from the positive class uniformly at random to make the number of samples in the positive class be the same as that in the negative class. This preprocessing is necessary for the training and testing of the CNN [36]. Then, for each of the two subreddits, we use 72% of uniformly randomly selected data (i.e., post titles) as training data, 8% as validation data, and 20% as the test data. To input post titles of different lengths to the CNN, we set the length of the input in terms of the number of words to the largest one among all the post titles, which was equal to 45 words after data cleaning. When an input post title is shorter than this length, we padded 0s after the post title to make the total length 45 words. Note that almost all the tweet samples (i.e., 99.94%) that we collected from Twitter, which we use as input to the trained CNN in the following analyses, are shorter in length than the maximum input length allowed for the CNN (i.e., 45 words after the cleaning). For any tweets longer than 45 words after the cleaning (i.e., at most 54 words after cleaning), we fed the first 45 words to the CNN. We employed Keras (https://keras.io/) using Tensorflow (https://github.com/tensorflow/tensorflow) as a backend to set up the neural network structure and train it.

The trained CNN is a softmax classifier, which outputs a value between 0 and 1. The output value is the ACE mention score value (See Section 2.3 for the ACE mention score) and represents the probability that the input text contains references to an ACE. If the output is above 0.5, the classifier judges the input to be associated with an ACE, which is part of the information used for training the CNN.

The training of the CNN also required a word embedding matrix. Word embeddings can capture the semantic meaning of words by converting them into numeric vectors [37] and for this we used Global Vectors for Word Representation (GloVe) [38]. GloVe is widely-used mapping from words to vectors, equivalent to a word embedding matrix whose rows and columns correspond to the words and the vector’s components, respectively.

To evaluate the classification performance of the trained CNN on the test data, we calculated the following five quantities [39]. We denote by TP the number of true positives (i.e., r/raisedbynarcissists posts that are correctly classified into the ACE-positive group), by TN the number of true negatives (i.e., r/internetparents posts that are correctly classified into the negative group), by FP the number of false positives (i.e., r/internetparents posts that are incorrectly classified into the positive group), and by FN the number of false negatives (i.e., r/raisedbynarcissists posts that are incorrectly classified into the negative group). The accuracy is equal to the fraction of the correct prediction, i.e., (TP+TN)/(TP+TN+FP+FN). The precision is given by TP/(TP+FP). The recall is given by TP/(TP+FN). The F1 score is the harmonic mean of the precision and recall, i.e., 2 × (Precision) × (Recall)/(Precision + Recall). Finally, we measured the area under the receiver operating characteristic curve (AUC). The receiver operating characteristic curve is the trajectory of the FP and TP, with FP on the horizontal axis and TP on the vertical axis, when we gradually increase the threshold for classification in terms of the output value of the CNN from 0 to 1. Note that we have fixed the threshold to 0.5 to actually classify the Reddit post titles and calculate the accuracy, precision, recall, and F1 score. Therefore, the definition of the FP and TP for calculating the AUC is different from that for calculating these four measures. A large AUC value indicates a good performance of binary classification.

### 2.3 ACE mention score for tweets and ACE alignment index for Twitter users

We used the Twitter Intelligence Tool (TWINT; https://github.com/twintproject/twint) to collect publicly available tweets, excluding retweets. We queried the tweets via the keywords explained in Section 2.5 (also see Table 1). The time frame used for the collection of the tweets was the same as that for the Reddit posts, i.e., from December 25, 2020 to March 31, 2022. We restricted ourselves to English tweets and otherwise did not use other filters. We then used the previously described classifier to score tweets in Twitter containing more than five words after cleaning (see Section 2.1 for the cleaning procedure). We refer to the computed output as the ACE mention score of the tweet, and it represents the probability that the tweet content contains references to an ACE.

**Table 1:**
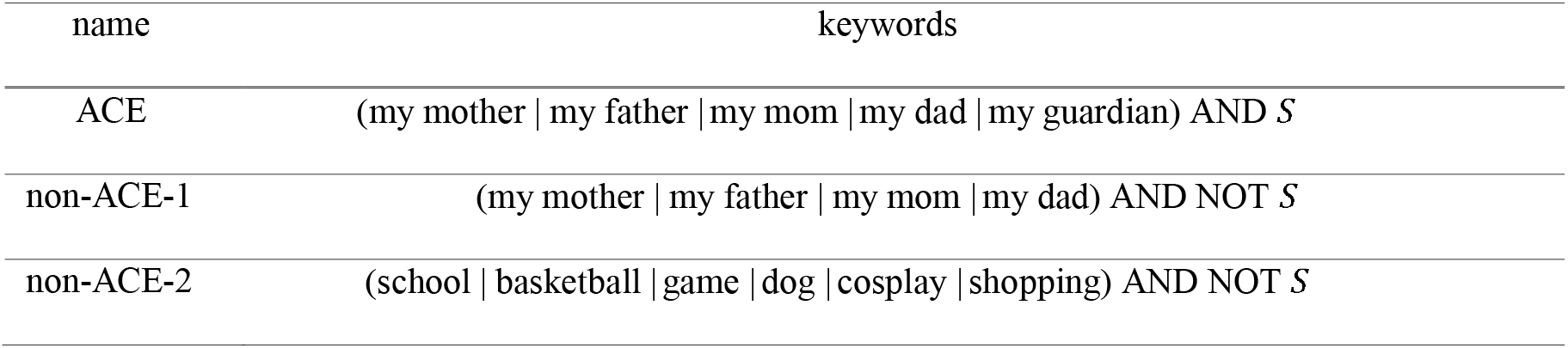
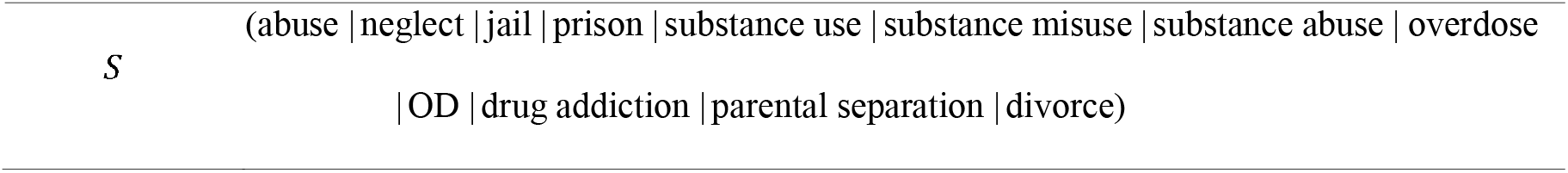
Keyword lists for sampling Twitter users. The vertical bars between words represents OR. OD abbreviates overdose.

Then, for each Twitter user that is the author of any of the collected tweets and has at least 30 tweets with more than five words after cleaning, we submitted each tweet posted by to the CNN classifier, obtaining its ACE mention score value. Then, we define the top 10% percentile value of the ACE mention score calculated from all tweets of as ‘s ACE alignment index. The intuition behind this definition is that individuals reporting ACEs would tweet about ACEs at least 10% of the time. The ACE alignment index ranges between 0 and 1 because the ACE mention score of each tweet ranges between 0 and 1.

If ‘s ACE alignment index is more than or equal to 0.5, we say that is an ACE individual. We manually inspected the ACE alignment index of the sampled Twitter users and their tweets to conclude that, while the threshold value of 0.5 for defining the ACE individual is reasonable, labeling all the individuals whose ACE alignment index is less than 0.5 non-ACE is inappropriate because there are many equivocal cases. Therefore, we define non-ACE users to be those whose all tweets with more than five words after the cleaning have ACE mention score less than 0.3. We show in Fig. 1 the entire process of calculating ‘s ACE alignment index and classifying into the ACE or non-ACE category.

**Figure 1:**
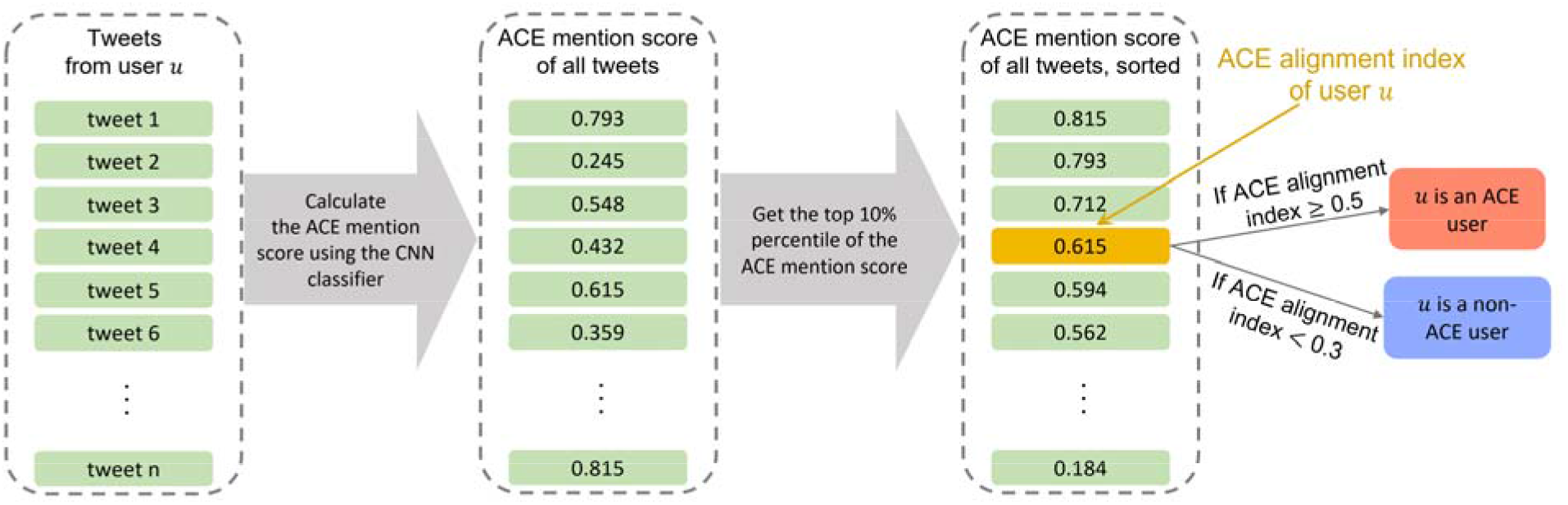
Schematic showing calculation of the ACE alignment index of a Twitter user.

### 2.4 Sentiment analysis

We calculated a standard sentiment score using a pretrained rule-based analysis model called VADER [40] to determine the sentiment of all tweets with more than five words after the cleaning posted by each Twitter user *u*. The sentiment score, denoted by *s*, ranges between −1 and 1, and *s* :5 −0.05, −0.05 < *s* < 0.05, and *s* > 0.05 indicate negative, neutral, and positive sentiment, respectively.

### 2.5 Egocentric networks of ACE and non-ACE Twitter users

We generated egocentric follow networks for ACE and non-ACE individuals. Nodes of a follow network are Twitter users. Each edge represents a following relationship, is directed from the follower to the followee, and is unweighted, as schematically shown in Fig. 2(a). We first needed to sample users from each category (i.e., ACE and non-ACE) whose egocentric networks we built. We call these users root users. To obtain root users, we ran a keyword search on all public tweets. With the aim of sampling tweets related to ACEs, we used the keyword list labeled “ACE” in Table 1. Notably, we added “my” before each word related to parent or guardian. This is because, without “my”, we obtained a large fraction of institutional and individual accounts that tweeted about ACEs but they themselves did not have ACEs [20]. With “my”, we intended to sample users who had self-reported ACEs. Then, we filtered the sampled users according to the following criteria. First, we examined public Twitter profiles and tweets to remove institutional and individual accounts that were advocating or supporting ACEs but had not tweeted about personal experiences. Second, for the ACE category, we obtained root users, for which the ACE alignment index is at least 0.5 from the steps described in Section 2.3.

**Figure 2:**
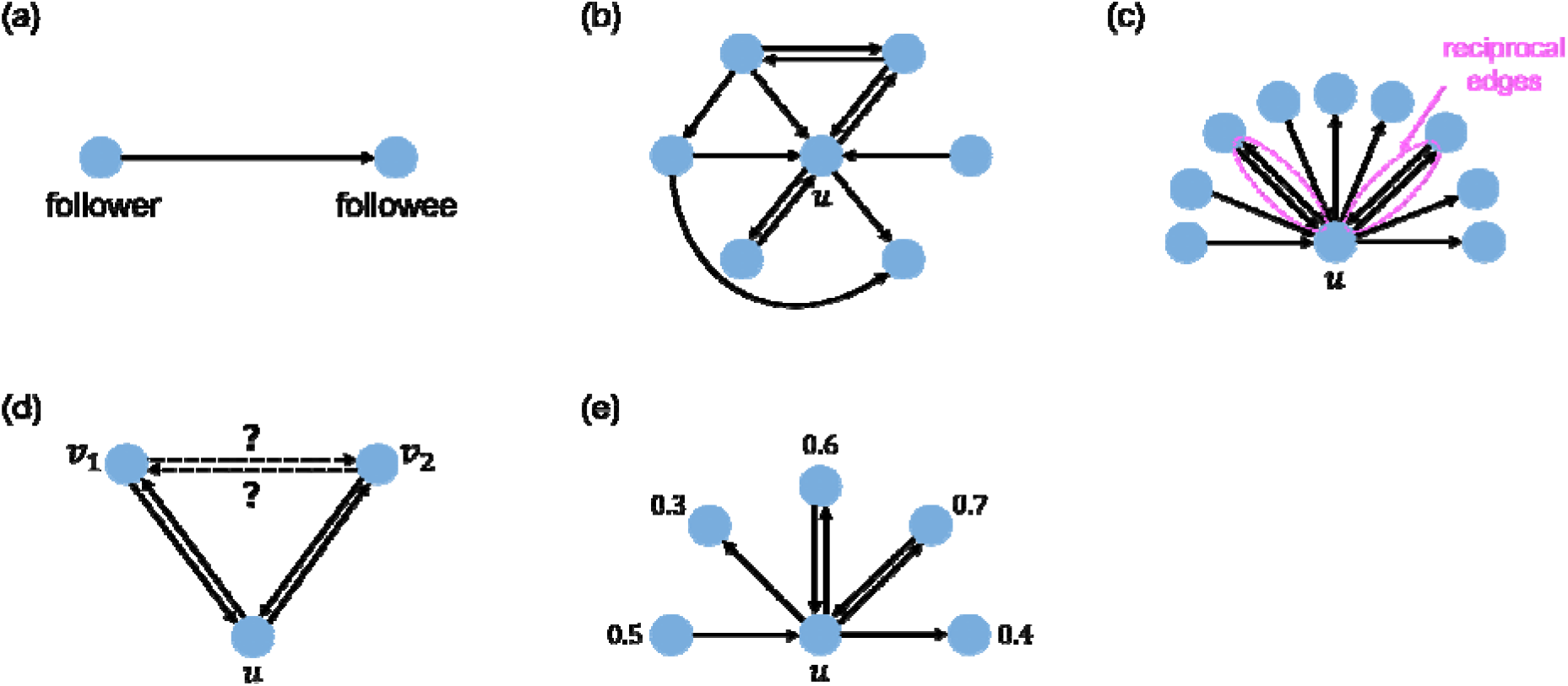
Schematic of the follow network and indices. (a) A directed edge, which points from a follower to a followee by definition. (b) A hypothetical example of egocentric network of a root user, . (c) Reciprocity indices. Root user has followers and followees. Because it has two reciprocal neighbors, one obtains and (d) Local clustering coefficients. Users and are reciprocal neighbors of . If and are reciprocal neighbors of each other, the reciprocal edge contributes to local clustering coefficients or as well as to or . If and are only unidirectionally connected, i.e., from to or vice versa but not both, the unidirectional edge contributes to or but not to or . (e) Calculation of the average ACE alignment index over the followers or followees. The numbers shown are the ACE alignment index. User has three followers and . User has four followees and.

To construct diverse samples of non-ACE root users for comparison purposes, we used each of the two keyword lists, named non-ACE-1 and non-ACE-2, shown in Table 1. The non-ACE-1 keyword list is informed by the ACE keyword list. To construct non-ACE1, we removed keywords related to events associated with ACE, such as “jail”, from the ACE keyword list. We also removed “my guardian” from the ACE keyword list to generate non-ACE-1 because “my guardian” suggests that sampled individuals have been separated from their biological parents and undergone foster care placement, which itself is an ACE. The non-ACE-2 keyword list is an arbitrarily chosen set of keywords that do not have to do with parenting. We filtered the users sampled with either non-ACE-1 or non-ACE-2 keyword list in the same manner as the case of the sampling of ACE individuals with the ACE keyword list, except that we retained the users whose all tweets had an ACE mention score less than 0.3. We refer to the final sets of root users obtained with these two keyword lists as non-ACE-1 and non-ACE-2 individuals.

For each root user *u* that belongs to either the ACE, non-ACE-1, or non-ACE-2 group, we collected their followers and followees (i.e., users that a user follows, which Twitter officially calls “following”) using the Python library Tweepy (https://github.com/tweepy/tweepy). We also collected the follow edges between *u*’s followers and followees if they existed. Due to the rate limit of the Twitter API, we only sampled up to 100 followers and 100 followees of each root user, and part of connectivity between pairs of them as we describe in Section 2.6. These connectivity data define egocentric networks of the root users (see Fig. 2(b) for an example; *u* represents a root user).

### 2.6 Network indices

Apart from the number of followers and followees, we measured the following three quantities for the egocentric network of each root user.

#### 2.6.1 Reciprocity

In directed networks, reciprocal edges, i.e., bidirectional edges, are considered to represent stronger relationships than unidirectional edges [41, 42]. In the case of the follow relation in Twitter, a reciprocal edge represents reciprocal following between two individuals (see Fig. 2(c)). Such pairs of individuals may be friends of each other. To compare the reciprocity of follow edges between ACE and non-ACE individuals, we measured two reciprocity indices for individuals defined as follows. For a given root user *u*, we sample *u*’s 100 followers uniformly at random. If *u* has less than 100 followers, we sample *u*’s all followers. In either case, we denote by *m*_1_ the number of the sampled followers. Then, among the *m* followers of *u*, we count the number of those who *u* follows back. We refer to such individuals as reciprocal neighbors of *u*. In other words, *u* both follows and is followed by each of its reciprocal neighbors. In the example shown in Fig. 2(c), the root user has *m*_1_ =5 followers and two reciprocal edges. Then, we define reciprocity *r*_1_ as the number of the reciprocal neighbors of *u* divided by *m*_1_. The root user shown in Fig. 2(c) has *r*_1_ = 2/5. We only sampled up to 100 followers due to the rate limit of the Twitter API. We avoided calculating *r*_1_when *u* had less than five followers because the calculated *r*_1_ value is considered to be unreliable when *m*_*1*_ is small.

Similarly, we sampled 100 followees of *u* selected uniformly at random or all the followees if *u* has less than 100 followees. We denote by *m*_*2*_ the number of the sampled followees. Reciprocity *r*_2_ is equal to the number of the reciprocal neighbors of *u* divided by *m*_*2*_. The root user shown in Fig. 2(c) has *m*_2_ =6 followees and therefore *r*_2_ = 2/6 = 1/3. We avoided calculating *r*_2_ when *u* had less than five followees. Thus, *r*_1_ is a reciprocity measure among a root user’s followers, and *r*_2_ is a reciprocity measure among a root user’s followees.

Both *r*_1_ and *r*_2_ range between 0 and 1. A unified measure of reciprocity for individual *u* would be the number of reciprocal edges divided by the number of any edges owned by *u* [42]. However, its computation requires collecting all the followers and followees, which is impossible due to the rate limit of the Twitter API. Therefore, we instead measured *r*_1_ and *r*_2_ for each root user.

#### 2.6.2 Clustering coefficients

The presence of triangles around an individual suggests that belongs to a group of at least three individuals, and such groups may provide social support to [25, 28, 43, 44]. Therefore, we measure the abundance of triangles around each root user by the sample local clustering coefficients defined as follows. Consider a root user . Then, we obtain the subset of the followers of that follows back. This subset defines a set of reciprocal neighbors of *u*. Then, we consider two reciprocal neighbors of *u* in this set, denoted by *v*_1_ and *v*_*2*_, and ask whether *v*_1_ and *v*_2_ are reciprocal neighbors of each other (see Fig. 2(d)). If they are, i.e., if *v*_1_ follows *v*_2_ and *v*_2_ follows *v*_1_, then *u, v*_1_ and *v*_*2*_ form a triangle in which each pair of individuals is connected by reciprocal edges. We define *u*’s local clustering coefficient, denoted by *C*_1_, as the fraction of (*v*_1_, *v*_2_) pairs that are reciprocal neighbors. We also measure a weaker version of the local clustering coefficient, denoted by 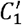, which only requires that *v*_1_ and *v*_1_ is adjacent by one follower edge in either direction (i.e., either *v*_1_ follows *v*_2_ or vice versa). We restricted *v*_1_ and *v*_2_ to be *u*’s reciprocal neighbors, not just followers or followees, because the clustering coefficient is primarily used for undirected networks. We avoided calculating *C*_1_ and 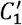 when *u* had less than five followers. Note that 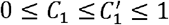.

We repeated the same measurements using a different set of reciprocal neighbors of *u*, which was the subset of the *m*_*2*_ followees of *u* that followed back *u*. We denote the thus calculated local clustering coefficients, depending on whether *v*_1_ and *v*_2_ are reciprocally connected or at least unidirectionally connected, by *C*_*2*_ and 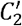 respectively. We avoided calculating *C*_*2*_ and 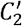 when *u* had less than five followees.

#### 2.6.3 Homophily

Lastly, we hypothesize that ACE individuals tend to be adjacent to other ACE individuals, presenting homophily. To test this hypothesis, we measured the fraction of ACE neighbors for ACE root users and non-ACE root users. To this end, for each root user *u*, we use its *m*_1_ followers sampled for the calculation of the reciprocity and local clustering coefficients. We calculated the ACE alignment index of each follower whose ACE alignment index can be calculated (i.e., those with at least 30 tweets with more than five words after the cleaning) and took the average over all such followers. This average, denoted by ⟨*α*⟩_follower_, ranges between 0 and 1 and defines the average ACE alignment index of *u*’s followers. In the example shown in Fig. 2(e), the root user *u* has three followers and ⟨*α*⟩_follower_ = 0.5 + 0.6 + 0.7 /3 = 0.6. We avoided calculating ⟨*α*⟩_follower_ for the root users *u* when we could not calculate the ACE alignment index for any of *u*’s followers, i.e., when none of *u*’s followers had at least 30 eligible tweets. We also measured the average ACE alignment index of *u*’s followees, denoted by ⟨*α*⟩_followee_, in the same manner. For example, the root user *u* in Fig. 2(e) has four followees and ⟨*α*⟩_followee_ = 0.3 + 0.6 + 0.7 + 0.4 /4 = 0.5.

We also compared the average ACE alignment index of the reciprocal neighbors of root users *u* and that of non-reciprocal followers of *u*. For example, individual *u* shown in Fig. 2(e) has two reciprocal followers, and their average ACE alignment index is (0.6 + 0.7)/2 = 0.65. The same individual has just one non-reciprocal follower, whose (average) ACE alignment index is 0.5. If the former tends to be larger than the latter as in this example, then *u* tends to follow back other ACE individuals than non-ACE individuals.

## 3 Results

### 3.1 Accuracy of the convolutional neural network

Our CNN trained using the Reddit data had an average accuracy of 82.78%, precision of 86.32%, recall of 78.07%, F1 score of 81.99%, and AUC of 91.34%. Using the same Reddit data set, we also trained the bidirectional encoder representations from transformers (BERT), which is a machine learning technique specialized to NLP [45]. However, its performance on the Reddit data was worse than that for the CNN (accuracy: 80.46%, precision: 80.93%, recall: 79.50%, F1 score: 80.21%, AUC: 89.31%). Therefore, we use the CNN in the following analyses to classify tweets.

### 3.2 Sampling ACE and non-ACE Twitter users

We sampled Twitter users with either of the three lists of keywords shown in Table 1 with the aim of sampling ACE and non-ACE individuals. We show in Fig. 3 the distribution of the ACE alignment index for the three groups of individuals, i.e., those sampled with the ACE keyword list, those sampled with the non-ACE-1 keyword list, and those sampled with the non-ACE-2 keyword list. Note that individuals sampled with the ACE keyword list may be non-ACE and vice versa. As expected, we find that the ACE alignment index for the individuals sampled with the ACE keyword list tends to be larger than that sampled with either of the two non-ACE keyword lists (ACE vs non-ACE-1: *p* < 10^−15^ ; ACE vs non-ACE-2: *p* < 10^−15^ ; MannWhitney-Wilcoxon test, two-sided, Bonferroni corrected including the comparison between non-ACE-1 vs non-ACE-2; we use the same statistical test in the following group comparison analyses). In particular, most users sampled with the ACE keyword list (97.6%; 123 out of the 126 sampled users) have an ACE alignment index of at least 0.5 (avg ± std = 0.81 ± 0.12). Those users are ACE individuals by definition.

**Figure 3:**
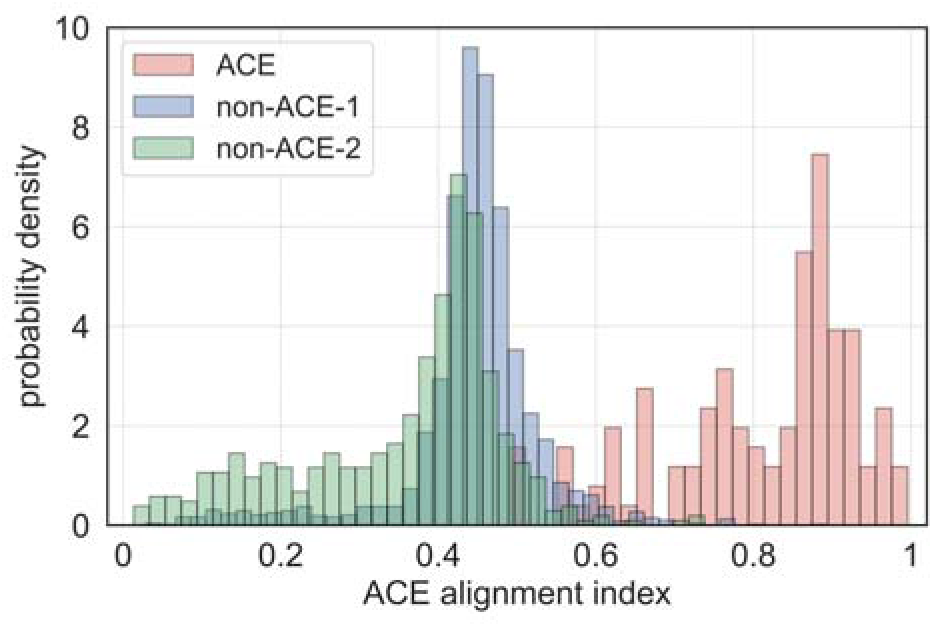
Distribution of the ACE alignment index for the three groups of Twitter users.

For validation, we also sampled Twitter users using a previously published list of keywords for ACEs [20]. With this keyword list, the ACE alignment index for the sampled individuals was 0.55 ± 0.10 (avg ± std), and 82% individuals had an ACE alignment index of at least 0.5. These numbers are substantially smaller than for the individuals sampled with our ACE keyword list (*p* < 10^−5^). Furthermore, the fraction of institutional accounts, which we needed to manually remove, was much larger with their ACE keyword list (13.7%) than with our ACE keyword list (2.4%). Therefore, we conclude that our ACE keyword list improves over the published one and continue to use the former in the following analyses.

Intriguingly, the ACE alignment index tended to be larger for the individuals sampled with the non-ACE1 keyword list than those sampled with the non-ACE-2 keyword list (avg ± std = 0.44 ± 0.09 for non-ACE-1 and 0.36 ± 0.13 for non-ACE-2; non-ACE-1 vs non-ACE-2: *p* < 10^−5^). In fact, 16.6% of the individuals sampled with the non-ACE-1 keyword list were ACE individuals (i.e., ACE alignment index ≥ 0.5), while a substantially smaller fraction of the individuals sampled with the non-ACE-2 keyword list (6.6%) was ACE individuals. This result indicates that individuals who tweet about their own parents tend to talk about ACEs more than those who do not. Mainly for this reason, we needed to sample many Twitter users with the non-ACE-1 keyword list to be able to sample non-ACE individuals, which are defined to be those whose maximum ACE mention score in their tweets is less than 0.3. We identified 119 (out of 2482) and 106 (out of 515) non-ACE individuals with the non-ACE-1 and non-ACE-2 keyword lists, respectively.

### 3.3 Sentiment of tweets

We analyzed sentiments of the tweets posted by the ACE, non-ACE-1, and non-ACE-2 root users. We show the distributions of the percentage of positive tweets and that of negative tweets for each of the three groups of root users in Fig. 4. We found that ACE individuals posted negative tweets more frequently than non-ACE individuals and that there was no significant difference in the fraction of positive tweets they posted.

**Figure 4:**
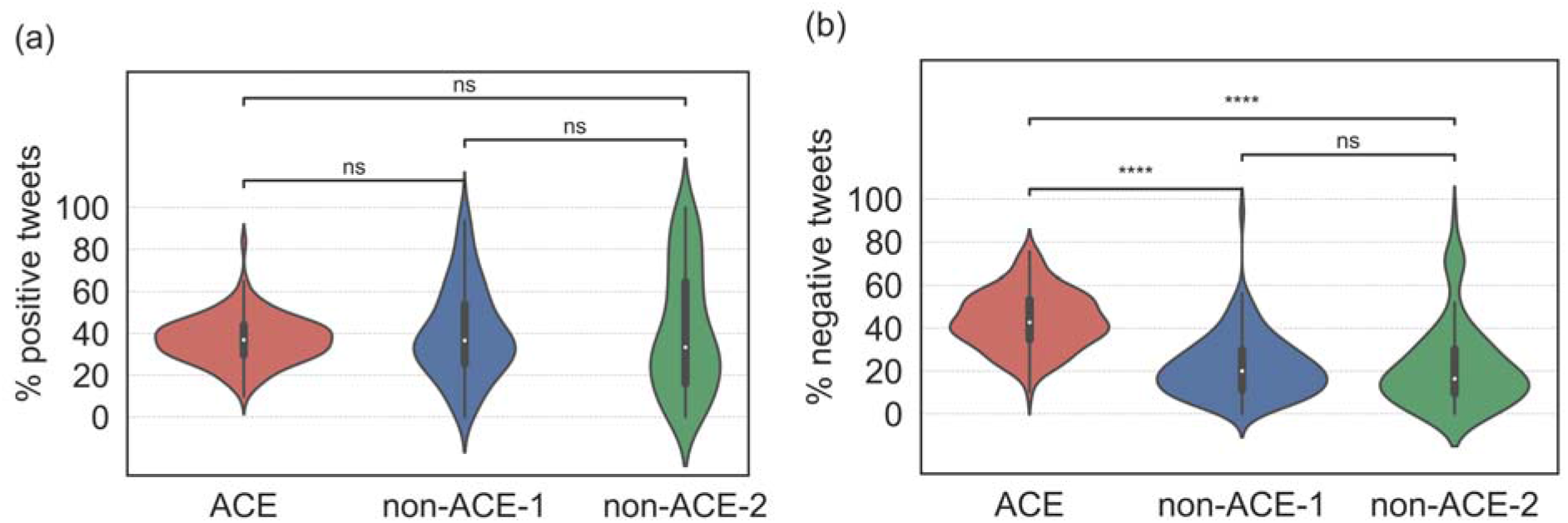
Distribution of the percentage of positive and negative tweets for ACE and non-ACE individuals. (a) Percentage of positive tweets. (b) Percentage of negative tweets. We used the violin plots implemented in Seaborn, a Python library, to visualize the distributions. The open circle in each violin plot represents the median. The thick vertical lines represent the interquartile range. The thin vertical lines represent the range. ns: not significant, *: *p* < 0.05, **: *p* < 0.01, ***: *p* < 10^−^, ****: *p* < 10^−^, all based on the two-sided Mann-Whitney-Wilcoxon test with Bonferroni correction.

### 3.4 Analysis of egocentric networks of ACE and non-ACE individuals

#### 3.4.1 Number of followers and followees

For the egocentric networks of the root users, we first investigated the number of followers and that of followees for each type of root users. We show the survival probability of the number of followers, *k*^in^, i.e., the fraction of the root users of which the number of followers is at least *k*^in^, in Fig. 5(a). Each distribution has a heavy tail, which is typical for distributions of the number of followers or followees in Twitter [46]. In other words, a small fraction of individuals has disproportionately many followers or followees compared to the majority. The number of followers was only significantly different between the ACE and non-ACE-1 groups (ACE vs non-ACE-1: *p* = 0.028; ACE vs non-ACE-2: *p* = 0.20; non-ACE-1 vs non-ACE-2: *p* = 0.93), with *k*^in^smaller for the ACE than the non-ACE-1 group. However, the non-significance result in the comparison of the ACE and non-ACE-2 groups is presumably due to the large standard deviations. In fact, their average *k*^in^ values were substantially different from each other (avg ± std = 433.8 ± 1058.9, min *k*^in^ = 0, max *k*^in^ = 5046 for ACE; avg ± std = 795.2 ± 1413.9, min *k*^in^ = 0, max *k*^in^ = 7089 for nonACE-1; avg ± std = 764.3 ± 1335.6, min *k*^in^= 0, max *k*^in^ = 6977 for non-ACE-2), Overall, ACE individuals tended to have fewer followers than non-ACE individuals, which is also notable in Fig. 5(a). This result may be because ACE individuals’ tweets attract less people than the tweets by non-ACE individuals on average. In contrast, the distribution of the number of followees, *k*^out^, was similar among the three groups (see Fig. 5(b); avg ± std = 551.6 ± 993.9, min *k*^out^ = 1, max *k*^out^ = 5213 for ACE; avg ± std = 687.6 ± 1080.3, min *k*^out^ = 0, max *k*^out^ = 5035 for non-ACE-1; avg ± std = 605 ±1039.7, min *k*^out^ = 0, max *k*^out^ = 5000 for non-ACE-2; ACE vs non-ACE-1: *p* = 0.93; ACE vs non-ACE-2: *p* = 0.39; non-ACE-1 vs non-ACE-2: *p* = 0.69).

**Figure 5:**
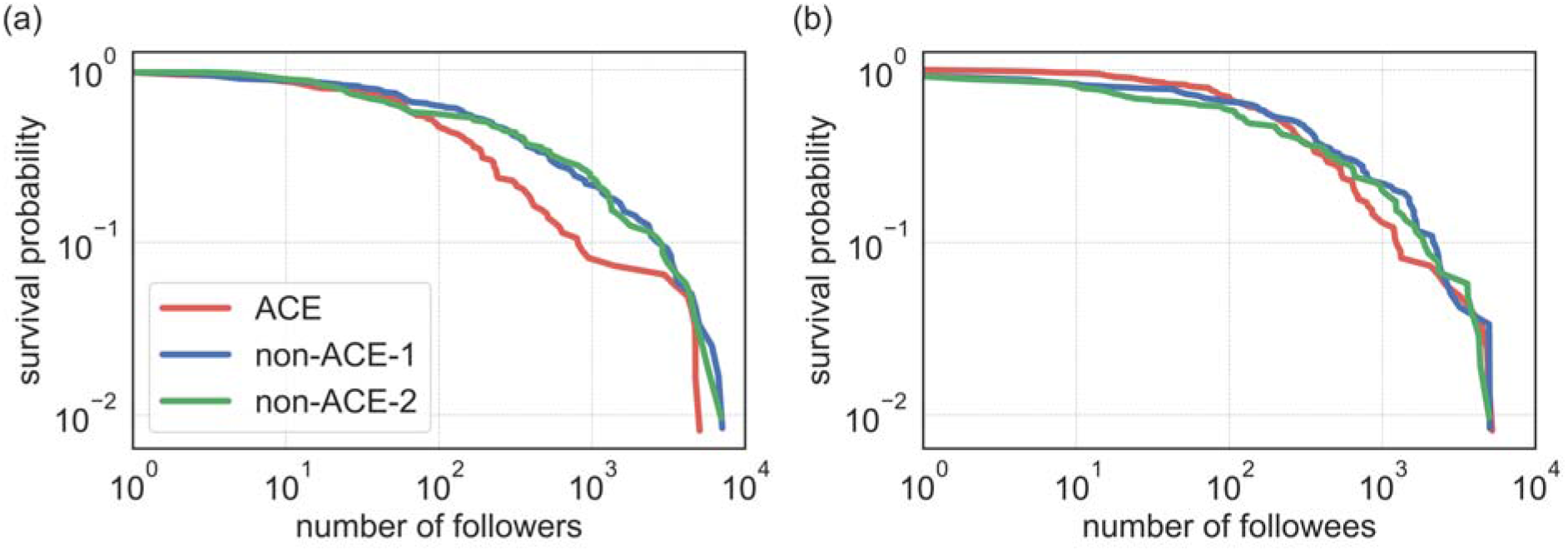
Survival probability of the distribution of the number of follow edges. (a) Number of followers. (b) Number of followees.

#### 3.4.2 Reciprocity

Next, to inspect possible differences in the structure of the egocentric network between ACE and non-ACE individuals, we investigated the reciprocity (i.e., fraction of mutual following between two individuals), local clustering coefficient (i.e., abundance of triangles), and homophily (i.e., fraction of ACE users in the immediate neighborhood of an ACE or non-ACE individual) for the root users.

We show the distribution of two types of reciprocity, *r*_1_ and *r*_2_, separately for the three groups of root users, as violin plots in Fig. 6. The *r*_1_ value was significantly different among the three groups. Specifically, for the ACE group was larger than that for the non-ACE-1 group, which was larger than that for the non-ACE-2 group. We did not find significant differences among the three groups in terms of *r*_2_. These results indicate that ACE individuals tend to follow back their followers, no matter which individual first started to follow the other individual, and that the individuals followed by a root ACE user do not particularly tend to follow back the root ACE user. Therefore, ACE individuals may tend to proactively connect to other individuals to yield reciprocal follow edges.

**Figure 6:**
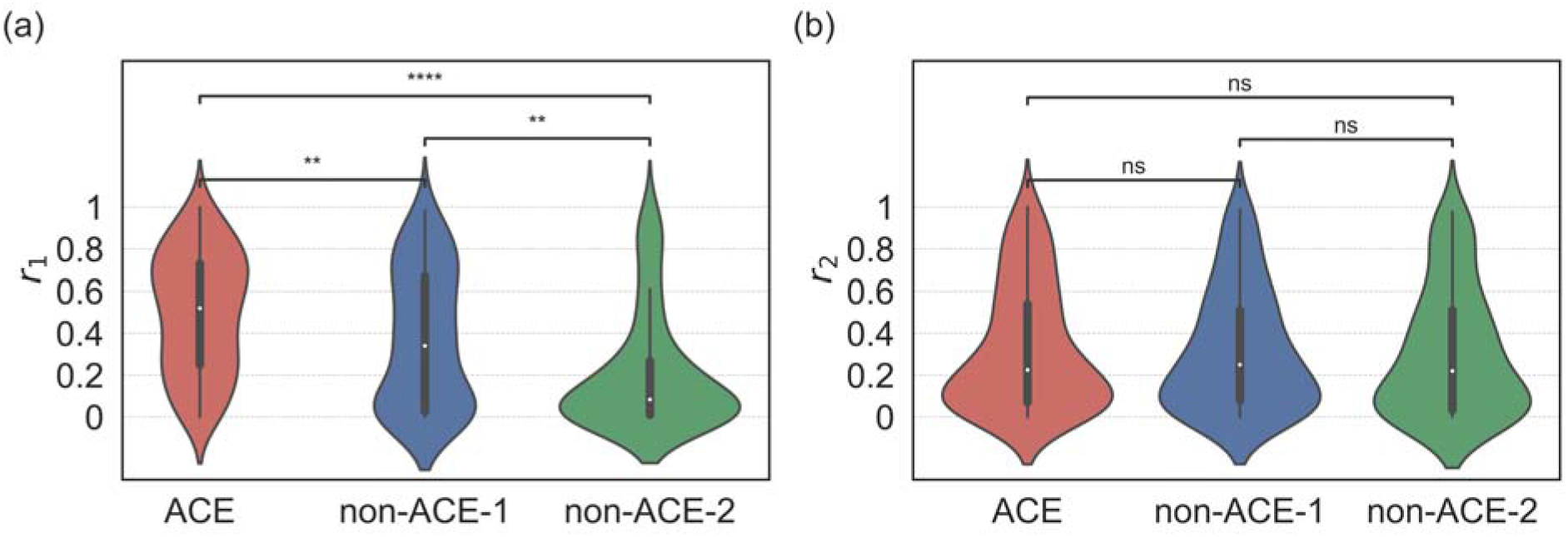
Distribution of the reciprocity for ACE and non-ACE individuals. (a) *r*_1_. (b) *r*_2_.

#### 3.4.3 Clustering coefficients

We show the distributions of the local clustering coefficients for the ACE and non-ACE groups in Fig. 7. The computation of the local clustering coefficient requires sampling of reciprocal neighbors of the root users *u*. When we collected *u* ‘s reciprocal neighbors by examining whether *u* followed back its followers, we only found significant results between the ACE and non-ACE-2 groups for the stronger and weaker definitions of the local clustering coefficient, i.e., *C*_1_ and 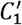, as shown in Fig. 7(a) and 7(b), respectively. In particular, there was no difference between the ACE and non-ACE-1 groups. When we collected *u*’s reciprocal neighbors by examining whether *u*’s follower followed *u* back, we did not find significant results between any pair of groups, either for the strong or weaker definitions of the local clustering coefficient (i.e., *C*_2_ and 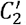; see Fig. 7(c) and 7(d), respectively). Overall, we conclude that the abundance of triangles around the root users, as quantified by the local clustering coefficients, is not different among the three groups in most cases.

**Figure 7:**
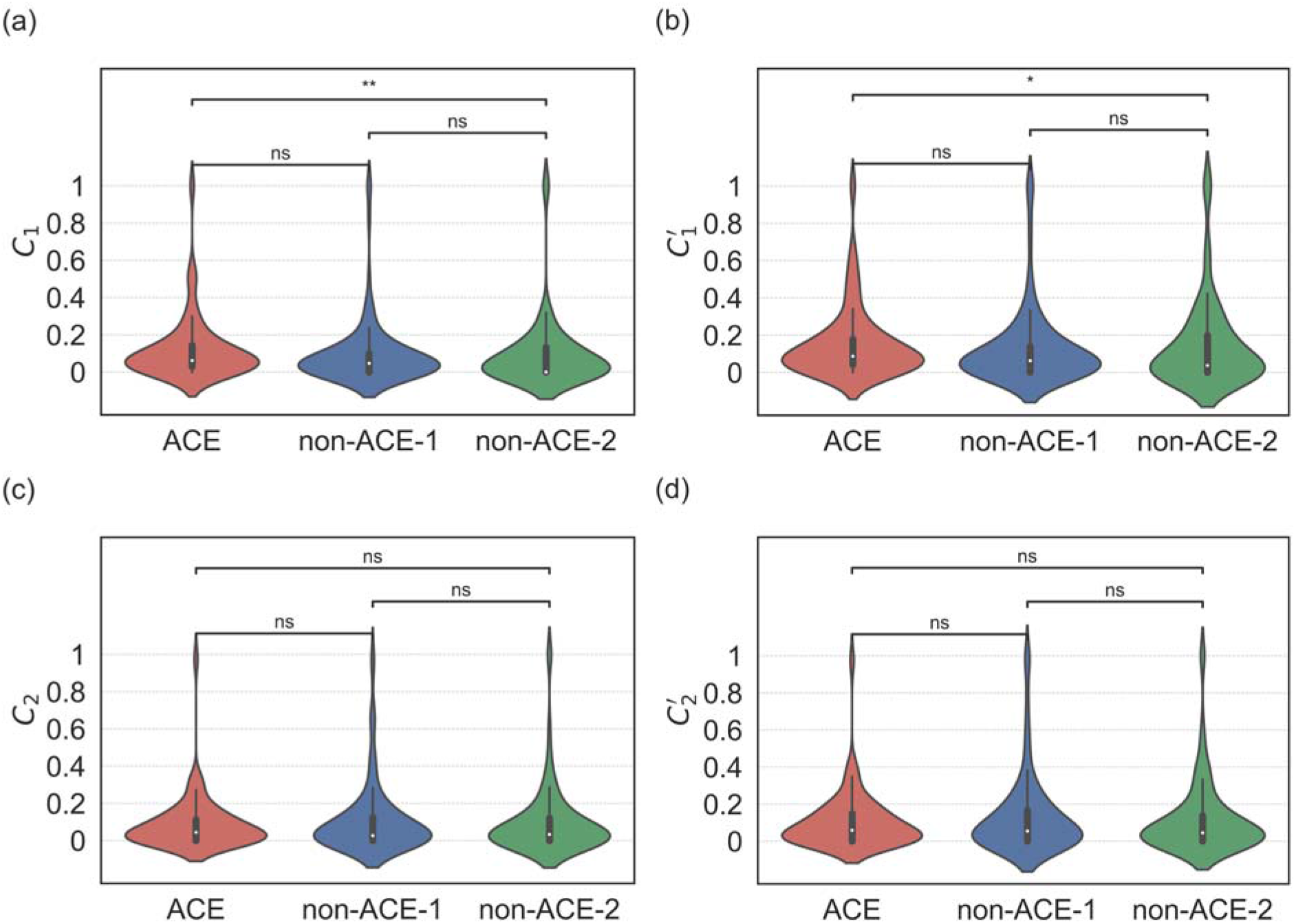
Distribution of the local clustering coefficient for ACE and non-ACE individuals. (a) *C*_1_. (b) 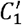 (c) *C*_2_. (d) 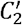.

#### 3.4.4 Homophily

The homophily in terms of ACEs would be a tendency that ACE individuals are preferentially connected to other ACE individuals in the network. To investigate the possibility of such homophily, we measured the average ACE alignment index of the followers, denoted by ⟨*α*⟩_follower_, for each ACE and non-ACE root user. We show the distribution of ⟨*α*⟩_follower_ for the different groups of root users in Fig. 8(a). The ACE alignment index for the followers of ACE individuals was significantly larger than that for the followers of non-ACE individuals, supporting the homophily hypothesis. We also found that the non-ACE-1 individuals had significantly larger ⟨*α*⟩_follower_ than the non-ACE-2 individuals. We then measured the average ACE alignment index of the followees, denoted by ⟨*α*⟩_followee_, for each root user. The distributions of ⟨*α*⟩_followee_ for the ACE, non-ACE-1, and non-ACE-2 groups of root users, shown in Fig. 8(b), were similar to those of ⟨*α*⟩_followee_ shown in Fig. 8(a), including the statistical results.

**Figure 8:**
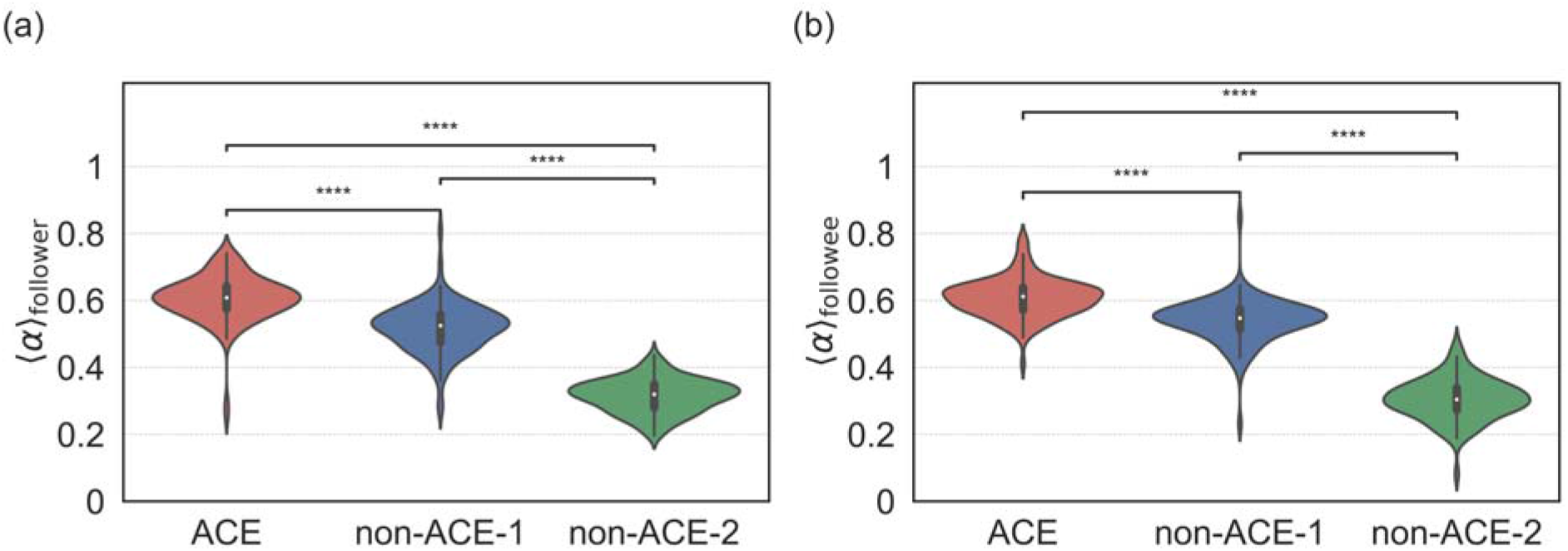
Distribution of the average ACE alignment index over the followers and followees of the ACE and non-ACE root users. (a) Average ACE alignment index over the followers, ⟨*α*⟩_follower_. (b) Average ACE alignment index over the followees, ⟨*α*⟩_followee_.

Figure 6 supports that ACE individuals tend to follow back their followers. Figure 8 supports that ACE individuals as compared to non-ACE individuals are more likely to be directly connected to other ACE individuals. The combination of these two results suggest that ACE individuals are more likely to follow back their ACE followers than their non-ACE followers. Therefore, we conducted a sub-analysis to compare the average ACE alignment index of the reciprocal neighbors of root users *u* and that of non-reciprocal followers of *u*.

We show the distribution of the average ACE alignment index of the reciprocal neighbors and that of the non-reciprocal followers in Fig. 9(a), (b), and (c) for ACE, non-ACE-1, and non-ACE-2 root users, respectively. The results statistically support that the ACE and non-ACE-1 root users, in particular the ACE root users, tended to follow back ACE individuals than non-ACE individuals, whereas the difference in the average ACE alignment index between the reciprocal neighbors and non-reciprocal followers was small. The difference between the reciprocal neighbors and non-reciprocal followers was not significant for the non-ACE-2 root users. Note that the present result that non-ACE-1 root users behaves more similarly to ACE root users than non-ACE-2 root users do is consistent with our results for the reciprocity (see Fig. 6), clustering coefficient (see Fig. 7(a) and (b)), and homophily (see Fig. 8).

**Figure 9:**
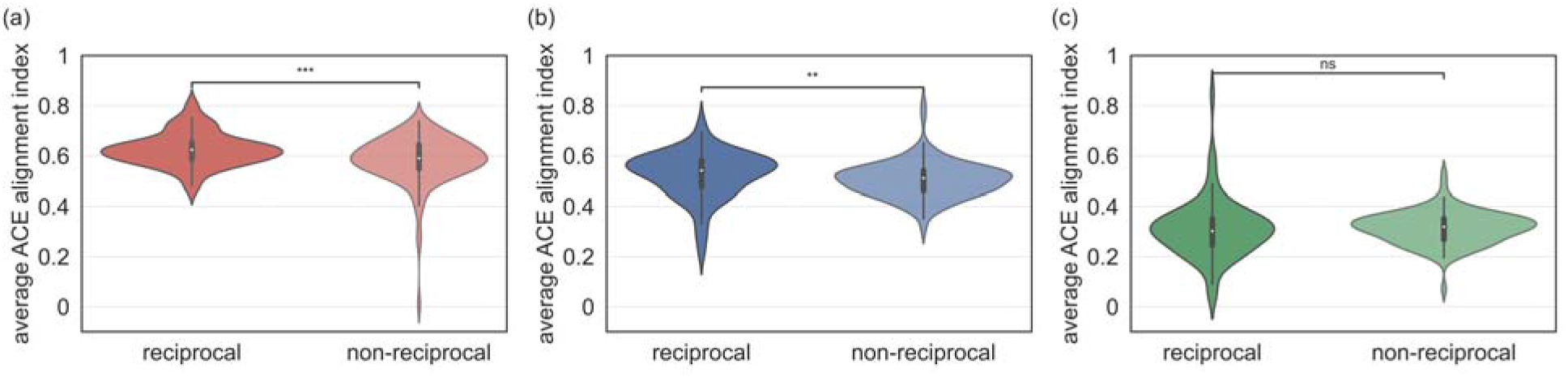
Distribution of the average ACE alignment index over the reciprocal neighbors and that over the non-reciprocal followers. (a) ACE root users. (b) Non-ACE-1 root users. (c) Non-ACE-2 root users.

## 4 Discussion

We trained a CNN model with Reddit post title data to classify tweets in Twitter into those associated with ACEs and those not. Using the trained CNN as a main tool, we sampled Twitter users, determined their strength of association with ACEs in terms of our ACE alignment index, and investigated the structure of egocentric follow networks of individuals reporting ACEs and those not reporting ACEs. We found that individuals reporting ACEs compared to non-ACE reporting individuals have fewer followers, similar numbers of followees (i.e., other individuals that an individual follows), a higher propensity to follow back, similar abundance of triangles around the individual (i.e., similar clustering coefficient values), more ACE individuals as their followers and followees, and a higher tendency to follow back other individuals reporting ACEs rather than other individual not reporting ACEs. Social networks of individuals reporting ACEs have been largely unknown except the number and quality of their immediate contacts. The present study significantly expands our understanding of their social networks by combining machine learning techniques, modern network analysis, and online social media data.

Studies suggest that connectedness through social networks can support individuals who have experienced ACEs [16, 17] or various mental health challenges [47] and confer a protective effect by disrupting the toxic stress pathways that connect adversity and trauma in childhood to poor health outcomes across the lifespan [19, 48]. The connectedness and social support that may emerge from social networks can also provide a positive adaptation and coping strategy, thereby reducing the need for maladaptive health risk behaviors to cope with the impact of ACEs. These studies and our present study share the main goal of revealing characteristics about the networks around individuals reporting ACEs. In one study [16], the authors quantified the social networks by the number of other individuals and social groups to which the participants belonged. In contrast, in addition to the number of directly connected other individuals, we examined the association between ACEs and further structural properties of the individuals’ social networks, i.e., the reciprocity of edges, local abundance of triangles, and homophily (i.e., the relative frequency with which ACE individuals follow other ACE individuals). In particular, we found that individuals reporting ACEs tend to make reciprocal follows and that they do so more frequently with other ACE individuals. This result may indicate that ACE individuals are actively seeking social support by connecting to other ACE individuals to share their ACEs and current lives. Existing strategies to prevent and respond to ACEs in trauma-informed ways take place in clinical, family, school, community, and some institutional settings [2]. Our results suggest the possibility of an additional venue – social networks and social media platforms – for prevention and mitigation strategies. This possibility has the added benefit of being more universally available, though attention to rural communities is in order, and accessible, particularly for those reluctant to seek support in more traditional and visible ways. Investigating the efficacy of this method warrants future work.

We found that Twitter users who mention their parents using the word “my” (e.g., “my mother”) tend to be associated with ACEs more likely than those who do not do so. This result is supported by the different distributions of the ACE alignment index, reciprocity, and homophily between the non-ACE-1 and non-ACE2 groups. We cannot rule out the possibility that this result is solely because users in the non-ACE-1 group tend to talk about their own parents and may lead to more misclassification of their tweets. However, this information may be useful for detecting and documenting ACEs in people’s online posts and questionnaire correspondences. Furthermore, systematically investigating whether people tend to use those words (e.g., “my father”) with negative meanings and sentiments when they use them, in particular on social media, is an intriguing research question.

Previous research showed that individuals with major depression [25, 26], suicide ideation [25, 28], or anxiety disorder [29] tended to have smaller local clustering coefficients (i.e., less triangles around the individuals) than healthy controls. Because triangles in networks are positively associated with social support [28, 43, 44], these results may indicate that the affected individuals are lacking in social support. In contrast, a different study using Twitter data showed that depressed individuals tended to have a higher local clustering coefficient than controls, suggesting that depressed individuals may prefer to build a closed network in which they want to share their experiences and obtain social supports [22]. Differently from these studies, we did not find differences in the local clustering coefficients between the ACE reporting and non-ACE reporting individuals. The reason for this result is unclear. However, it does not contradict our interpretation of the main results of the present study that the ACE individuals’ high reciprocity and homophily in the following behavior may reflect social support that they seek (i.e., connection to other ACE individuals). Investigating the nature of triangles, such as who are in the triangle, requires more data, such as exhaustive sampling of the followers and followees of the root users. However, these tasks are computationally difficult due to the rate limit imposed by Twitter. Future work employing different social media and other types of data containing information about triangles may be able to better understand this topic.

We used Reddit and Twitter data to sample and assess individuals reporting ACEs. There are at least two strengths of this approach compared to conventional questionnaire-based methods. First, the employed data are observational. Because Reddit or Twitter users’ posting behavior is unrestricted by experimenters, the obtained data are expected to be less subject to recall and other biases, which data collection based on retrospective reporting is generally subject to [49-51]. A second strength is scalability. Although the free Twitter API is rate-limited, one can still collect information about tweets and follow relationships of hundreds and thousands of individuals without much difficulty. In contrast, a potential main drawback of social media data is credibility. For example, a previous study pointed out that credibility and quality of tweet data about ACEs are not guaranteed [20]. Another clear limitation of Twitter and some other social media data is that data are poorly annotated. For example, our results may be confounded by nuisance parameters such as the individual’s age, sex, education, geographical location, economic status, and race. Many people do not reveal such information on Twitter, and there is no personal information on users’ public profiles on Reddit. Therefore, part of the significant differences among the three groups revealed in the present study may be due to unmeasurable factors such as different demographic characteristics of the three groups. This limitation makes the present study difficult to define control groups, and thus we employed two non-ACE groups (i.e., non-ACE-1 and non-ACE-2). The lack of annotation also makes it difficult to estimate the severity of ACEs, such as the standard ACE score [52] for Twitter users. There are clear trade-offs between the advantages of social media data and of questionnaire-generated data. Validating the present results with clinical populations, such as by asking them about specific usages of Twitter or other social media data, may benefit both social-media-based and questionnaire-based studies of ACEs.

There are also other limitations of the present study. First, we used the Twitter’s follow network as a proxy to social interaction between individuals. However, follow behavior, including the case of reciprocal following, does not necessarily represent a friendship or reasonable social relationship [46, 53]. Future studies should look into other social media networks or combine social media data and questionnaire data to better estimate social networks including the meaning of the edge (i.e., link) of the network.

Second, there may be transferability issues between the Reddit and Twitter data. In fact, the accuracy of the CNN on the Twitter data was high as long as we sampled some ACE and non-ACE Twitter users and manually inspected their tweets. However, users on Reddit, which our CNN was trained on, may be statistically different from Twitter users in terms of demography. It is also likely that people using both Reddit and Twitter often publish Reddit posts and tweets with different intentions and in different situations. Although transfer learning is a common technique in data science, it is desirable to enhance homogeneity of the two populations by better sampling and user profiling.

Third, there is no established approach to ACE-related search terms on Twitter data. We defined our search terms via a combination of literature [20] and expert’s knowledge in our team. However, public discourse and social media users may not use the same language to refer to childhood trauma and adversity as experts. Therefore, we may have missed individuals reporting ACEs in Reddit and Twitter using their own terms. Thorough text analyses of posts in online ACE communities are expected to help better definitions of search terms for similar studies in the future.

Fourth, one may be able to improve the accuracy of our CNN using a different type of classifier, or a larger amount of training data including those from different subreddits and other social media platforms relevant to ACEs. In fact, our CNN is not specialized to NLP, and we showed that the BERT, a classifier specialized to NLP, did not improve the classification performance (see Section 3.1). Despite these limitations, we believe that the present study clarifies new features of social networks with people with ACEs and suggests opportunities of future research on ACEs involving social media data, network analysis, and machine learning techniques.

## Data Availability

All data produced in the present study are available upon reasonable request to the authors.

https://github.com/ycao20/ACE-project/

## Acknowledgements

The authors acknowledge Freelancer Technology Pty Limited, in particular, Ihsan Ali, Tiffany Fong, Anna Lazzereschi, and Joshua Tan for running the data science open competition leading to this work and managing the collaboration among the authors after the open competition.

## References

[1] V. J. Felitti, R. F. Anda, D. Nordenberg, D. F. Williamson, A. M. Spitz, V. Edwards, M. P. Koss, and J. S. Marks. Relationship of childhood abuse and household dysfunction to many of the leading causes of death in adults: The Adverse Childhood Experiences (ACE) study. Am. J. Prev. Med., 14:245–258, 1998

[2] Centers for Disease Control and Prevention. Preventing Adverse Childhood Experiences: Leveraging the Best Available Evidence. Atlanta, GA: National Center for Injury Prevention and Control, Centers for Disease Control and Prevention, 2019

[3] L Bynum and et al. Adverse childhood experiences reported by adults—Five states, 2009. MMWR Morbidity Mortality Weekly Report, 59:1609–1613, 2010.

[4] J. S. Carlson, J. Yohannan, C. L. Darr, M. R. Turley, N. A. Larez, and M. M. Perfect. Prevalence of adverse childhood experiences in school-aged youth: A systematic review (1990–2015). Int. J. School Edu. Psychol., 8:2–23, 2020.

[5] Centers for Disease Control and Prevention. Adverse Childhood Experiences (ACEs): Preventing early trauma to improve adult health. Atlanta, GA: National Center for Injury Prevention and Control, Centers for Disease Control and Prevention, 2019

[6] S. M. Brown, J. R. Doom, S. Lechuga-Peña, S. E. Watamura, and T. Koppels. Stress and parenting during the global COVID-19 pandemic. Child Abuse Negl., 110:104699, 2020.

[7] D. J. Bryant, M. Oo, and A. J. Damian. The rise of adverse childhood experiences during the COVID-19 pandemic. Psychol. Trauma, 12:S193–S194, 2020.

[8] J. Cuartas. Heightened risk of child maltreatment amid the COVID-19 pandemic can exacerbate mental health problems for the next generation. Psychol. Trauma, 12:S195–S196, 2020.

[9] D. M. Sperry and C. S. Widom. Child abuse and neglect, social support, and psychopathology in adulthood: A prospective investigation. Child Abuse Negl., 37:415–425, 2013.

[10] J. M. Horan and C. S. Widom. From childhood maltreatment to allostatic load in adulthood: The role of social support. Child Maltreatment, 20:229–239, 2015.

[11] S. Lagdon, J. Ross, M. Robinson, A. A. Contractor, R. Charak, and C. Armour. Assessing the mediating role of social support in childhood maltreatment and psychopathology among college students in Northern Ireland. J. Interp. Viol., 36:NP2112–NP2136, 2021.

[12] R. Schwarzer and A. Leppin. Social support and health: A theoretical and empirical overview. J. Soc. Pers. Rel., 8:99–127, 1991.

[13] B. N. Uchino. Social support and health: A review of physiological processes potentially underlying links to disease outcomes. J. Behav. Med., 29:377–387, 2006.

[14] L. F. Berkman and T. Glass. Social integration, social networks, social support, and health. In L. F. Berkman and I. Kawachi, editors, Social Epidemiology, pages 137–173. Oxford University Press, Oxford, UK, 2000.

[15] C. A. Heaney and B. A. Israel. Social networks and social support. In K. Glanz, B. K. Rimer, and K. Viswanath, editors, Health Hehavior and Health Education: Theory, Research, and Practice, 4:189–210. John Wiley & Sons, Inc., San Francisco, CA, fourth edition, 2008.

[16] M. McLafferty, J. Ross, B. Waterhouse-Bradley, and C. Armour. Childhood adversities and psychopathology among military veterans in the US: The mediating role of social networks. J. Anxiety Disorders, 65:47–55, 2019.

[17] F. D. Schneider, C. A. Loveland Cook, J. Salas, J. Scherrer, I. N. Cleveland, and S. K. Burge. Childhood trauma, social networks, and the mental health of adult survivors. J. Interp. Viol., 35:1492–1514, 2020.

[18] N. A. Christakis and J. H. Fowler. Connected. Little, Brown and Company, New York, NY, 2009.

[19] D. Easley and J. Kleinberg. Networks, Crowds, and Markets. Cambridge University Press, Cambridge, UK, 2010.

[20] A. Srivastav, K. Park, A. Koziarski, M. Strompolis, and J. Purtle. Who is talking about adverse childhood experiences? Evidence from Twitter to inform health promotion. Health Edu. Behav., 48:615–626, 2021.

[21] M. A. Moreno, L. A. Jelenchick, K. G. Egan, E. Cox, H. Young, K. E. Gannon, and T. Becker. Feeling bad on Facebook: Depression disclosures by college students on a social networking site. Depression and Anxiety, 28:447–455, 2011.

[22] M. De Choudhury, M. Gamon, S. Counts, and E. Horvitz. Predicting depression via social media. In: Proc. 7th International Conference on Weblogs and Social Media, ICWSM 2013, 2:128–137, 2013.

[23] S. C. Guntuku, R. Schneider, A. Pelullo, J. Young, V. Wong, L. Ungar, D. Polsky, K. G. Volpp, and R. Merchant. Studying expressions of loneliness in individuals using twitter: an observational study. BMJ Open, 9:e030355, 2019.

[24] K. Saha, A. Yousuf, R. L. Boyd, J. W. Pennebaker, and M. De Choudhury. Social media discussions predict mental health consultations on college campuses. Sci. Rep., 12:123, 2022.

[25] N. Masuda, I. Kurahashi, and H. Onari. Suicide ideation of individuals in online social networks. PLoS ONE, 8:e62262, 2013.

[26] S. Negriff. Depressive symptoms predict characteristics of online social networks. J. Adol. Health, 65:101– 106, 2019.

[27] J. A. Lam, B. Lu, N. J. Doogan, T. Thomson, A. Ferketich, E. D. Paskett, and M. E. Wewers. Depression, smoking, and ego-centric social network characteristics in Ohio Appalachian women. J. Rural Mental Health, 41:30–41, 2017.

[28] P. S. Bearman and J. Moody. Suicide and friendships among American adolescents. Am. J. Public Health, 94:89–95, 2004.

[29] S. Dutta and M. De Choudhury. Characterizing anxiety disorders with online social and interactional networks. Lect. Notes Comput. Sci., 12427:249–264, 2020.

[30] A. Degnan, K. Berry, D. Sweet, K. Abel, N. Crossley, and D. Edge. Social networks and symptomatic and functional outcomes in schizophrenia: a systematic review and meta-analysis. Soc. Psychiatry Psychiatric Epid., 53:873–888, 2018.

[31] S. Chancellor, G. Nitzburg, A. Hu, F. Zampieri, and M. De Choudhury. Discovering alternative treatments for opioid use recovery using social media. In Proc. 2019 CHI conference on human factors in computing systems, pages: 1–15, 2019.

[32] M. Hussain, J. J. Bird, and D. R. Faria. A study on cnn transfer learning for image classification. In UK Workshop on Computational Intelligence, pages 191–202, Cham, Switzerland, 2018. Springer.

[33] Y. Luan and S. Lin. Research on text classification based on CNN and LSTM. In Proc. 2019 IEEE International Conference on Artificial Intelligence and Computer Applications, ICAICA 2019, pages 352– 355, 2019.

[34] G. Gkotsis, A. Oellrich, S. Velupillai, M. Liakata, T. J. P. Hubbard, R. J. B. Dobson, and R. Dutta. Characterisation of mental health conditions in social media using Informed Deep Learning. Sci. Rep., 7:45141, 2017.

[35] J. Kim, J. Lee, E. Park, and J. Han. A deep learning model for detecting mental illness from user content on social media. Sci. Rep., 10:11846, 2020.

[36] V. López, A. Fernández, S. García, V. Palade, and F. Herrera. An insight into classification with imbalanced data: Empirical results and current trends on using data intrinsic characteristics. Info. Sci., 250:113–141, 2013.

[37] F. K. Khattak, S. Jeblee, C. Pou-Prom, M. Abdalla, C. Meaney, and F. Rudzicz. A survey of word embeddings for clinical text. J. Biomed. Info., 4:100057, 2019.

[38] J. Pennington, R. Socher, and C. D. Manning. GloVe: Global vectors for word representation. In Proc. 2014 Conf. on Empirical Methods in Natural Language Processing (EMNLP), pages 1532–1543, 2014.

[39] D. M. W. Powers. Evaluation: From precision, recall and F-measure to ROC, informedness, markedness & correlation. J. Mach. Learn. Technol., 2:37–63, 2011.

[40] C. J. Hutto and E. Gilbert. VADER: A parsimonious rule-based model for sentiment analysis of social media text. In Eighth Intl. AAAI Conf. Weblogs and Social Media, 8:216–225, 2014.

[41] S. Wasserman and K. Faust. Social Network Analysis. Cambridge University Press, New York, NY, 1994.

[42] D. Garlaschelli and M. I. Loffredo. Patterns of link reciprocity in directed networks. Phys. Rev. Lett., 93:268701, 2004.

[43] D. Krackhardt. The ties that torture: Simmelian tie analysis in organizations. Research in the Sociology of Organizations, 16:183–210, 1999.

[44] G. Viry. Residential mobility and the spatial dispersion of personal networks: Effects on social support. Soc. Netw., 34:59–72, 2012.

[45] J. Devlin, M.-W. Chang, K. Lee, and K. Toutanova. BERT: Pre-training of deep bidirectional transformers for language understanding. Preprint arXiv:1810.04805, 2018.

[46] S. A. Myers, A. Sharma, P. Gupta, and J. Lin. Information network or social network? In Proc. 23rd International Conference on World Wide Web, pages 493–498, 2014.

[47] B. L. Perry and B. A. Pescosolido. Social network dynamics and biographical disruption: The case of “first-timers” with mental illness. Am. J. Sociol., 118:134–175, 2012.

[48] J. P. Shonkoff, A. S. Garner; Committee on Psychosocial Aspects of Child and Family Health; Committee on Early Childhood, Adoption, and Dependent Care; Section on Developmental and Behavioral Pediatrics. The lifelong effects of early childhood adversity and toxic stress. Pediatrics., 129(1): e232–46, 2012.

[49] R. E. Norman, M. Byambaa, R. De, A. Butchart, J. Scott, and T. Vos. The long-term health consequences of child physical abuse, emotional abuse, and neglect: A systematic review and meta-analysis. PLoS Med., 9:e1001349, 2012.

[50] K. A. Kalmakis and G. E. Chandler. Health consequences of adverse childhood experiences: A systematic review. J. Am. Assoc. Nurse Practitioners, 27:457–465, 2015.

[51] K. Hughes, M. A. Bellis, K. A. Hardcastle, D. Sethi, A. Butchart, C. Mikton, L. Jones, and M. P. Dunne. The effect of multiple adverse childhood experiences on health: a systematic review and meta-analysis. Lancet Publ. Health, 2:e356–e366, 2017.

[52] R. E. Lacey and H. Minnis. Practitioner review: Twenty years of research with adverse childhood experience scores — Advantages, disadvantages and applications to practice. J. Child Psychol. Psychiatry, 61:116–130, 2020.

[53] H. Kwak, C. Lee, H. Park, and S. Moon. What is twitter, a social network or a news media? In Proc. 19th International Conference on World Wide Web, pages 591–600. ACM, 2010.

